# Social support, social strain and declines in verbal memory: 16-year follow-up of the English Longitudinal Study of Ageing cohort

**DOI:** 10.1101/2021.05.29.21258037

**Authors:** Shaun Scholes, Jing Liao

## Abstract

**Objective:** Estimate differences in the rate of decline in verbal memory by levels of perceived relationship quality among community-dwelling adults.

**Participants:** In the English Longitudinal Study of Ageing (ELSA), n = 10,109 participants aged 50-89 years were assessed at wave 1 (baseline: 2002-03) and followed-up over 16 years to wave 9 (2017-18).

**Methods:** Verbal memory was assessed biennially by tests of immediate and delayed word-recall. Positive and negative aspects of perceived relationship quality (social support and strain, respectively) were measured by relationship type (spouse; children; extended family members; friends). Random effects within-between (REWB) modelling was used to separate between- and within-person effects. Associations were estimated between levels of social support/strain and (i) baseline levels of memory (main effects), and (ii) the 2-year decline in memory (interaction with time).

**Results:** Longitudinal associations were most prominent for men, specific to relationship type, and showed between-rather than within-person effects. Among men, higher spousal strain was associated with faster decline in memory (β_between-effect×time_ = -0.043; 95% CI: -0.084, -0.002; p = 0.039), whilst greater support from children was associated with slower decline (β_between-effect×time_ = 0.020; 95% CI: 0.002, 0.039; p = 0.033). Men with higher levels of strain from friends had lower baseline memory (β_between-effect_ = -0.382; 95% CI: -0.627, -0.137; p = 0.002) and showed faster decline (β_between-effect×time_ = -0.047; 95% CI: -0.095, 0.000; p = 0.051).

**Conclusions:** Differences between persons in levels of social support and social strain were modestly associated with the rate of memory decline, especially among men. Our findings can inform future research studies and intervention strategies designed to maximise the potential of social relations to promote healthy cognitive ageing.

## Introduction

Prospective studies of ageing UK cohorts have shown levels of cognitive task performance to be inversely associated with morbidity and mortality (1, 2). Identifying the modifiable risk factors for cognitive ageing can inform future interventions. Socially supportive relationships serve as a coping resource to protect individuals’ physical- and mental-health (3). The World Health Organization identifies social support as a key social environmental factor that can enhance healthy ageing (4); for older adults, social support - particularly from the spouse and children - represents the main source of informal care to improve quality of life (5) and protect against progression of functional limitations (6).

Social relations are complex and multidimensional (7). However, the greater specificity of their structural, functional and qualitative aspects has improved scientific understanding of how social relations influence health-related outcomes (8). First, the structural aspects capture the observable features of social networks (e.g. number of ties, network composition, and contact frequency). Second, the supportive or functional aspects capture the actual exchange of aid (e.g. instrumental support), affect (e.g. emotional support), and affirmation. Third, the qualitative aspects capture subjective evaluations of the quality of relationships (e.g. levels of satisfaction, enjoyment, strain or conflict) (9). As a potential modifiable risk factor, interest has grown in identifying the specific ways in which social relations can best promote healthy cognitive ageing (10). The structural aspects of social relations have been linked with age-related cognitive change through the mentally stimulating nature of social interactions (10, 11). Fewer studies have explored the qualitative aspects: yet, the number of people with whom one interacts may be less important than the perceived quality of relationships (9).

Identifying the pathways through which the qualitative aspects of social relations influence cognitive ageing requires separating positive and negative interactions (12). For example, Zahodne et al (2019) distinguish between high-quality positive relationships, characterised by social support, and negative-quality relationships, characterised by social strain (e.g. excessive demands, conflicts, tension and criticism) (10). Stress regulation is postulated to be the key mechanism through which these qualitative aspects influence cognitive ageing: with supportive relations being a coping resource to reduce perceived stress, whilst strained relations act as a stressor (10, 11).

According to the solidarity-conflict model, social support and strain are not competing or antagonistic concepts but can coexist among close relationships; understanding how these work together in tandem to influence health outcomes is therefore more informative than examining them in parallel (13). To date, mixed evidence has accumulated regarding whether positive (social support) and negative (social strain) social exchanges independently influence cognitive ageing. In the MacArthur Studies of Successful Aging, greater frequency of emotionally supportive interactions with a network of social relations assessed at baseline was associated with lower cognitive decline over a 7.5 year period, adjusting for levels of demands/criticism from network members (14). In the Midlife in the U.S (MIDUS) cohort, lower frequency of social strain/conflict (but not social support) predicted higher executive function a decade later; whilst the frequency of social support (but not strain/conflict) was positively associated with episodic memory (15). In the UK Whitehall II cohort, after adjustment for positive aspects, participants in the top third of reported cumulative negative aspects of close relationships (adverse social interactions producing worries, problems, and stress, and needing more support) experienced a faster 10-year decline in executive function than those in the bottom third (16).

However, a number of key gaps in scientific understanding remain. More work is therefore needed to refine understanding on the ways in which the qualitative aspects of social relations influence cognitive ageing. First, few studies have examined these associations separately by sex (17) or by relationship type (10, 18, 19). Men and women maintain social relationships differently, having different requirements and expectations (20). Women have larger, denser, and more diverse social networks than men (21), and women both benefit from and are burdened by providing and receiving support from multiple sources (22, 23). Men maintain close relationships with fewer people, primarily their spouse (24), and they receive most social support from intimate ties (21). Higher levels of spousal support/strain may therefore strongly associate with cognitive ageing more for men than for women.

Secondly, by measuring social support/strain only at baseline (10, 14) or cumulatively (i.e. social histories) (15, 16), previous studies have not fully investigated the within- and between-person associations between social support/strain and cognitive ageing. Within-person change in perceived relationship quality (e.g. relationships being more supportive/strained than usual) may influence cognitive ageing independently of differences between persons in levels of social support/strain. However, studies employing standard modelling techniques have not fully disentangled between-person variation from within-person variation in longitudinal associations by either: (i) neglect the role of differences between persons by focusing exclusively on within-person change (fixed effects models), or (ii) implicitly assume that the within- and between-person effects are equal (random effects/ mixed models) (25).

The advantages of fixed effects and random effects models are combined in ‘random effects within-between’ models (REWB, also known as hybrid models), which simultaneously model the within- and between-person effects of a single time-varying independent variable (26). Applying REWB models to study the social relationships/cognitive ageing associations may inform the design of different interventions. For example, counselling therapies may be effective in buffering any short-term impacts of social relations that are more strained than usual (addressing within-person effects); whilst initiatives to boost levels of engagement with social technology may increase the amount of social support for persons with lower levels relative to their peers (addressing between-person effects).

To improve understanding on whether the positive and negative aspects of relationship quality associate with cognitive ageing, the present authors published a study in 2017 based on 8-years follow-up of the English Longitudinal Study of Ageing (ELSA) cohort (waves 1 to 5: 2002-03 to 2009-10) (27). Using REWB models, we explored the sex-specific longitudinal associations between social support/strain and age-related cognitive change separately by relationship type (spouse; children; family; friends). In the present study, we update our work by including the 4 most recent cognitive assessments to extend the follow-up period to 16-years (waves 1 to 9: 2002-03 to 2018-19). Including more measurements enables investigation of longer-term changes in cognitive functioning (10); as described by Gottesman et al (2014), studies with longer versus shorter follow-up periods may be better placed to evaluate the most critical time period during which most cognitive decline occurs (28). Lengthening the follow-up period however potentially increases the risk of selection bias arising from missing cognition data due to study drop-out, including death. Therefore, we use joint modelling of longitudinal and survival data in a sensitivity analysis to examine the robustness of our main findings. After adjustment for socio-demographics and physical- and mental-health, we hypothesise that: (H1) higher levels of social support, and lower levels of social strain, associate independently with higher initial levels of cognition and with slower decline; (H2) associations show both within- and between-person effects; and (H3) associations differ by sex and by relationship type.

## Methods

### Study design and participants

ELSA is an ongoing study of community-dwelling adults in England; *n* = 11,391 persons (born before March 1, 1952) participated in wave 1 (67% response rate). A detailed description of the goals, design and methods of ELSA is available elsewhere (29). Data collection takes place biennially via face-to-face interviews in the participant’s home, and, after a computer-assisted personal interview, self-completion questionnaires are filled in. To minimise potential for reverse causality (30), we excluded participants at wave 1 with doctor-diagnosed Alzheimer or Parkinson disease, dementia, or serious memory impairment. Proxy respondents, those aged 90+, and those with missing baseline cognition data were also excluded. Participants provided signed consent for taking part in the study and for linkage to mortality data; ethical approval was granted by the London Multicentre Research Ethics Committee (MREC/01/2/91).

### Assessment of verbal memory

Verbal memory was our chosen outcome measure as tests were administered at each wave. Participants were presented with a list of 10 words that were read out by a computer at the rate of 1 word for every 2 seconds. Participants were then asked to recall as many words as they could (immediate recall); they were asked to recall these words after an interval during which they completed other cognitive tests (delayed recall). Both scores were summed to obtain a composite continuous measure of words correctly recalled (range, 0-20); these were normally distributed, suggesting the absence of floor or ceiling effects. Word-recall tests have shown good construct validity and consistency (31).

### Social support and social strain

Questions on social relationship quality were administered at each wave via self-completion, and covered four relationship types: (i) spouse/partner; (ii) children; (iii) extended family members; and (iv) friends. Three items examined the participants’ perception of social support (positive evaluations of relationship quality): (i) “*How much do they really understand the way you feel about things?*”; (ii) “*How much can you rely on them if you have a serious problem?*”; and (iii) “*How much can you open up to them if you need to talk about your worries?*”. These items cover empathy, dependability, and confiding, respectively. Responses ranged from “not-at-all” (scored 0) to “a lot” (scored 3). Scores from each relationship were calculated separately using the average of the 3 items (1 or 2 items in the case of item missingness). A global score was calculated by averaging the 4 scores. Participants without the relevant social ties were scored zero.

Three items examined negative evaluations of relationship quality (i.e. social strain): (i) “*How much do they criticize you?*”; (ii) “*How much do they let you down when you are counting on them?*”; and (iii) “*How much do they get on your nerves?*”. These items cover criticism, being let down, and annoyance, respectively. Responses were scored as described for social support (higher scores indicated higher strain).

### Confounders

Based on previous research (30, 32), we identified the following time-independent confounders (assessed at wave 1): age (range: 50-89 years) and socioeconomic position (SEP: wealth and education). Time-dependent confounders were healthy lifestyle behaviours (smoking, alcohol consumption and physical activity); social participation; physical functioning; and depressive symptoms. Total wealth represented the sum of financial, physical and housing wealth, minus debts, and was grouped into quintiles (lowest to highest). Education status was categorised as low (compulsory schooling); medium (up to high school) and high (university degree or higher). Smoking status was classified as current cigarette smoker or not; alcohol consumption was classified as ever drinking or not. Participants were asked how often they engaged in moderate and in vigorous sports/activities: we classified participants as physically active or inactive. A social participation score was created based on involvement in 8 activities related to civic participation, leisure activities and cultural engagement. For physical functioning, a mobility limitation score was created based on the number of reported difficulties in performing 6 basic activities of daily living tasks (ADL); a score for the number of reported depressive symptoms was created based on the 8-item Center for Epidemiologic Studies Depression Scale (33).

### Statistical analyses

All analyses were weighted using the wave 1 weight to ensure that the sample was broadly representative of the community-dwelling English population aged 50+ years at baseline. Means and standard deviations (SD) for continuous variables and percentages for categorical variables were calculated to present the sample characteristics by study wave. Linear random/mixed effects models with study-wave-since-baseline as timescale (range, 0-8) were used to estimate baseline levels and change (slope, per 2-year increase in follow-up time) in memory. We ran sex-specific models to examine potential differences in the longitudinal associations (H3). We conducted three model-based analyses.

#### Unadjusted model: decline in verbal memory by age at baseline

First, we estimated nonlinearity in the rate of memory decline by including linear and quadratic terms (i.e. time^2^); baseline age (centred at 65 years) and age^2^; and their statistical interaction (time × age) as predictors in the fixed (population-average) part of the model.

#### REWB models with global measures of social support/strain

Secondly, we added the global measures of social support/strain as independent variables in REWB models. A single time-varying independent variable is included twice by: (i) assigning the respective variable the same value over all waves (each participant’s mean score across all waves), and (ii) allowing each participant’s score to vary over time (by use of a deviation score: the difference between the wave-specific and mean value) (26). The former estimates between-person effects; the latter estimates within-person effects.

As social support and strain may independently influence cognitive ageing (H1; H2), the models contained four terms of primary interest: the main effects and their interaction with time (linear change only). For the main effects, the between-person effect (β_between-effect_) represents the difference in baseline memory per unit difference between participants in their mean level of social support/strain; the within-person effect (β_within-effect_) represents the (population-averaged) difference in baseline memory for a given participant whose level of social support/strain at baseline was 1 unit higher than their usual level (34). Terms for the interaction with time (e.g. β_between-effect×time_) represent the absolute difference in the linear slope per unit increase in social support/strain; positive and negative coefficients indicate that a unit increase in social support/strain slowed and increased the rate of memory decline, respectively.

The fully-adjusted model contained terms for time, time^2^, age, age^2^, social support, social strain, and the potential confounders listed above. Interaction terms for each were included to represent differences in the linear slope. We adjusted for the number of prior word-recall tests (range, 0-8) to correct for practice effects (35); this term also proxies characteristics that influence attrition, which in turn, strongly associate with cognitive task performance (36). Our analyses were restricted to participants who filled in the self-completion questionnaires to minimise the amount of item missingness on social support/strain. To increase statistical precision and power, we used the multiple imputation using chained equations (MICE) method (37). Estimates from the REWB models were combined across 10 imputed datasets using Rubin’s rules.

#### Models stratified by relationship type

In our third analysis, we fit models separately for each relationship type (H3).

#### Sensitivity analysis

We used joint modelling of the longitudinal outcome (memory scores) alongside a survival model for time to death (38) to assess the impact of biases associated with study drop-out on our main findings. This model assumes that the association between the survival and the longitudinal processes is underlined by shared random effects (39). Joint modelling is particularly useful for adjusting for informative drop-out due to death (e.g. if participants with lower memory scores have lower survival over the follow-up period than those with higher scores, resulting in nonignorable drop-out and a reduction in statistical power to detect the influence of social relations on a limited range of memory change) (39). The REWB model and the Weibull proportional hazards model were assumed for the longitudinal and survival submodels, respectively. Data was analysed using Stata v16.1 (Stata Corp LP, College Station, Texas). Statistical significance tests were based on two-sided probability (*p* < 0.05).

## Results

The analytical sample (memory data and filled in self-completion questionnaires) comprised *n* = 10,109 participants aged 50-89 at wave 1 (2002-03); whom contributed 49,286 observations over the 9 waves (Figure S1). The sample was unbalanced due to study drop-out: *n* = 1901 (19%) participants contributed data only at wave 1, whilst *n* = 2004 (20%) contributed data in all 9 waves (data not shown). On average, participants contributed 4.9 (SD 3.0) waves of data.

Table 1 shows the characteristics of the key variables for those remaining in the study over time. Mean age at wave 1 was 64 years, fewer than half were male (47%), and over one-third had completed no more than compulsory schooling (41%). At wave 1, participants correctly recalled on average 9.5 (SD 3.5) words. The global measures of social support and social strain showed similar levels over time.

**TABLE 1.** Analytical sample characteristics by study wave

| Characteristics | Wave 1<br>2002-03<br>Mean (SD) | Wave 2<br>2004-05<br>Mean (SD) | Wave 3<br>2006-07<br>Mean (SD) | Wave 4<br>2008-09<br>Mean (SD) | Wave 5<br>2010-11<br>Mean (SD) | Wave 6<br>2012-13<br>Mean (SD) | Wave 7<br>2014-15<br>Mean (SD) | Wave 8<br>2016-17<br>Mean (SD) | Wave 9<br>2018-19<br>Mean (SD) |
| --- | --- | --- | --- | --- | --- | --- | --- | --- | --- |
| Participants (n) | 10,109 | 7395 | 6184 | 5271 | 5197 | 4638 | 3996 | 3463 | 3033 |
| **Time-dependent:** |  |  |  |  |  |  |  |  |  |
| Age, years | 64 (10.1) | 66 (9.7) | 67 (9.4) | 69 (9.0) | 70 (8.0) | 71 (7.5) | 72 (7.0) | 73 (6.5) | 75 (6.6) |
| Verbal memory<sup>a</sup> | 9.5 (3.5) | 10.1 (3.4) | 10.2 (3.6) | 10.3 (3.5) | 10.3 (3.6) | 10.4 (3.6) | 10.2 (3.6) | 10.2 (3.7) | 10.1 (3.7) |
| Social support<sup>b</sup> | 1.9 (0.6) | 1.9 (0.6) | 2.0 (0.6) | 1.9 (0.6) | 1.9 (0.6) | 1.9 (0.6) | 1.9 (0.6) | 1.9 (0.6) | 1.9 (0.6) |
| Social strain<sup>b</sup> | 0.6 (0.4) | 0.6 (0.4) | 0.6 (0.4) | 0.5 (0.4) | 0.5 (0.4) | 0.5 (0.4) | 0.5 (0.4) | 0.5 (0.4) | 0.5 (0.4) |
| Social participation | 1.5 (1.5) | 1.6 (1.5) | 1.5 (1.4) | 1.5 (1.4) | 1.5 (1.4) | 1.6 (1.4) | 1.6 (1.5) | 1.6 (1.4) | 1.6 (1.4) |
| Depressive symptoms (CESD) | 1.5 (1.9) | 1.5 (1.9) | 1.4 (1.9) | 1.3 (1.8) | 1.4 (1.9) | 1.2 (1.7) | 1.3 (1.7) | 1.2 (1.7) | 1.3 (1.7) |
| Mobility limitations (ADL) | 0.4 (0.9) | 0.4 (0.9) | 0.4 (0.9) | 0.3 (0.8) | 0.3 (0.9) | 0.3 (0.9) | 0.3 (0.9) | 0.4 (0.9) | 0.4 (0.9) |
| Current smoker (%) | 18 | 14 | 13 | 12 | 11 | 9 | 9 | 7 | 6 |
| Ever drunk alcohol (%) | 89 | 89 | 89 | 88 | 87 | 85 | 85 | 85 | 84 |
| Physically inactive (%) | 17 | 15 | 15 | 16 | 18 | 18 | 18 | 21 | 22 |
| **Time-independent:** |  |  |  |  |  |  |  |  |  |
| Men (%) | 47 | 46 | 46 | 46 | 45 | 45 | 45 | 46 | 45 |
| Compulsory schooling (%) | 41 | 37 | 35 | 33 | 32 | 30 | 29 | 27 | 25 |
| Lowest wealth (%) | 19 | 17 | 16 | 15 | 15 | 15 | 15 | 14 | 14 |

ADL: activities of daily living; CESD: Center for Epidemiologic Studies Depression Scale; SD: standard deviation.
The number of participants (n) is shown for those with non-missing memory scores and who filled in the self-completion questionnaire: participants with missing data on the other variables are excluded from this table.
<sup>a</sup> Sum of correctly recalled words (immediate and delayed).
<sup>b</sup> Scores averaged across 4 sources: spouse; children; extended family; friends.

### Memory trajectories by age at baseline

Based on models unadjusted for social support/strain, memory scores initially remained fairly stable but then declined more rapidly per 2-year time interval (β_time_ ^2^: *p* < 0.001; Figure 1 and Table S1); reaching the maximum at 2.2 and 3.0 years-since-baseline for men and women aged 65, respectively.

**Figure 1.**
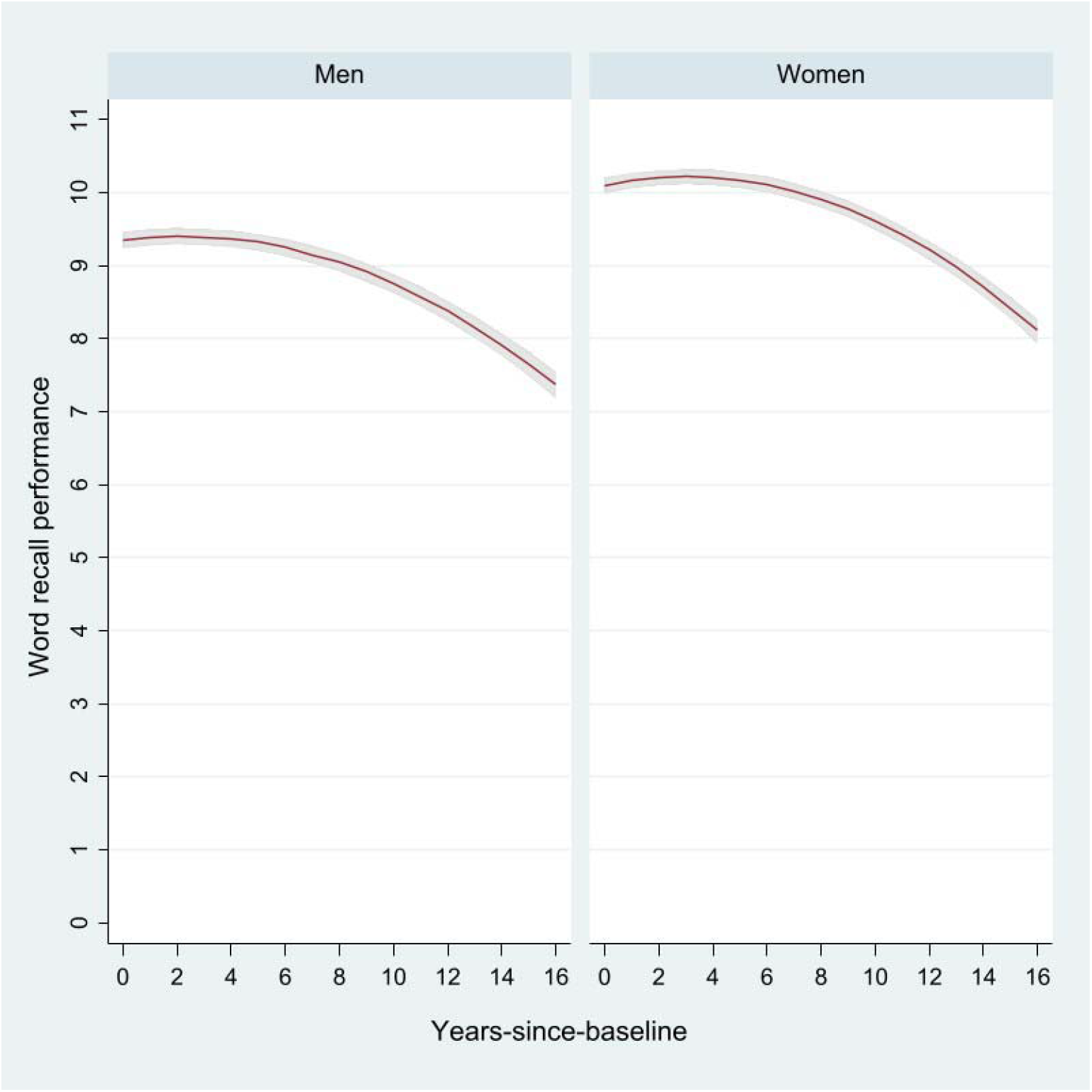
Verbal memory trajectories by sex

### REWB models with global levels of social support/strain

Adjusting for global levels of social support/strain, being younger, higher educated, higher wealth, having ever drunk alcohol, having fewer depressive symptoms, and being physically active were associated with higher initial memory for both sexes (β_main-effects_: all *p* < 0.003; Table S2). Slower memory decline was associated with younger age among both sexes (β_age×time_: *p* < 0.001). Though men in the lowest educational group showed lower baseline memory than those with a degree or higher qualification, their rate of decline in memory was lower (β_low-educ×time_: *p* = 0.005). Among women, slower memory decline was associated with being a non-smoker (β_smoke×time_: *p* = 0.006); higher social activity (β_social participation×time_: *p* = 0.030); and having fewer mobility limitations (β_ADL×time_: *p* = 0.025). These associations were similar to those found in previous investigations of cognitive trajectories in the ELSA cohort (30), and were similar in the models stratified by relationship type (data not shown).

Separately for social support/strain, the coefficients for the between- and within-person effects, and their interaction with time, after adjustment for socio-demographics and physical- and mental-health, are shown in Table 2.

**TABLE 2.** Results from REWB models of the between-persons and within-person associations between social support/strain and verbal memory (all sources)

| Type of association by sex | Social support |  |  | Social strain |  |  |
| --- | --- | --- | --- | --- | --- | --- |
| | $\beta$ | 95% CI | P-value | $\beta$ | 95% CI | P-value |
| *Men (n = 4595)* |  |  |  |  |  |  |
| Between-effect | 0.070 | -0.082, 0.221 | 0.367 | **-0.441** | **-0.703, -0.179** | **0.001** |
| Between-effect × time | 0.030 | -0.002, 0.061 | 0.067 | -0.029 | -0.083, 0.026 | 0.302 |
| Within-effect | -0.005 | -0.184, 0.174 | 0.954 | -0.069 | -0.296, 0.159 | 0.554 |
| Within-effect × time | -0.027 | -0.074, 0.021 | 0.269 | 0.004 | -0.058, 0.065 | 0.909 |
| *Women (n = 5514)* |  |  |  |  |  |  |
| Between-effect | 0.007 | -0.143, 0.158 | 0.925 | **-0.479** | **-0.732, -0.225** | **<0.001** |
| Between-effect × time | 0.021 | -0.010, 0.052 | 0.187 | 0.025 | -0.025, 0.075 | 0.331 |
| Within-effect | 0.056 | -0.123, 0.236 | 0.534 | 0.179 | -0.011, 0.369 | 0.065 |
| Within-effect × time | -0.025 | -0.069, 0.019 | 0.265 | -0.043 | -0.093, 0.006 | 0.084 |

$\beta$ : beta coefficient; CI: confidence interval; REWB: random effects within-between model

Higher mean levels of social strain were associated with lower baseline memory among men (β_between-effect_ = -0.441; 95% CI: -0.703, -0.179; *p* = 0.001) and among women (β_between-effect_ = -0.479; 95% CI: -0.732, -0.225; *p* < 0.001). Higher social support was marginally associated with a slower rate of memory decline among men (β_between-effect×time_ = 0.030; 95% CI: -0.002, 0.061; *p* = 0.067).

### Models stratified by relationship type

Results from the models for each relationship type are shown in Table 3.

**TABLE 3.**
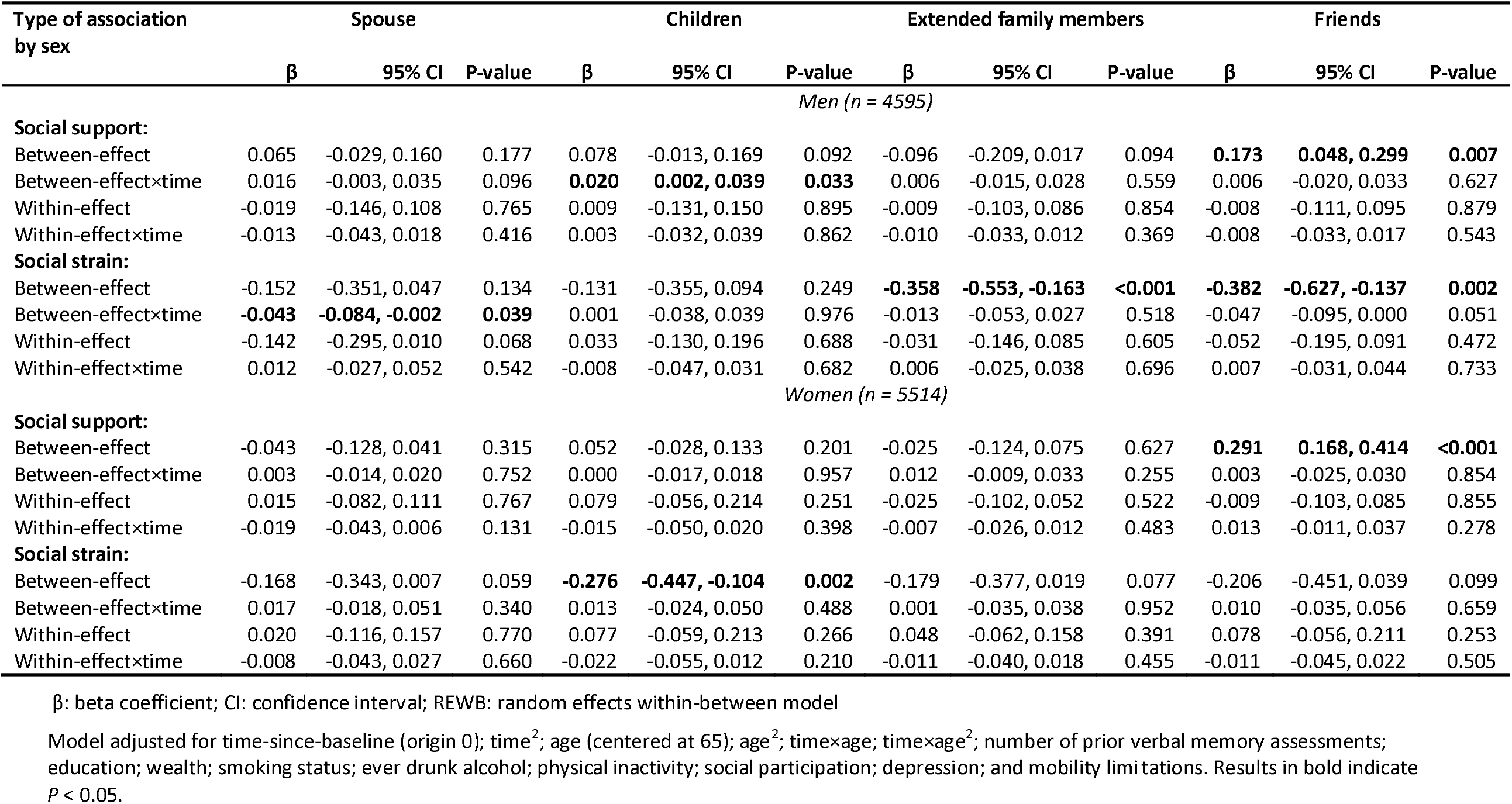
Results from REWB models of the between-persons and within-person associations between social support/strain and verbal memory by relationship type

#### Spouse

Faster decline in memory was observed among men with higher mean levels of spousal strain (β_between-effect×time_ = -0.043; 95% CI: -0.084, -0.002; *p* = 0.039). Among men, a higher-than-usual level of spousal strain was marginally associated with lower baseline memory (β_within-effect_ = -0.142; 95% CI: - 0.295, 0.010; *p* = 0.068). Women with higher mean levels of spousal strain had marginally lower baseline memory (β_between-effect_ = -0.168; 95% CI: -0.343, 0.007; *p* = 0.059).

#### Children

Slower decline in memory was observed among men with higher levels of support from children (β_between-effect×time_ = 0.020; 95% CI: 0.002, 0.039; *p* = 0.033). Women with higher mean levels of strain from children had lower baseline levels of memory (β_between-effect_ = -0.276; 95% CI: -0.447, -0.104; *p* = 0.002).

#### Extended family members

Men with higher mean levels of strain from extended family members had lower baseline memory (β_between-effect_ = -0.358; 95% CI: -0.553, -0.163; *p* < 0.001); a similar but weaker association was observed among women (β_between-effect_ = -0.179; 95% CI: -0.377, 0.019; *p* = 0.077).

#### Friends

Men with higher levels of social strain from friends had lower baseline memory (β_between-effect_ = -0.382; 95% CI: -0.627, -0.137; *p* = 0.002) and showed faster decline (β_between-effect×time_ = -0.047; 95% CI: -0.095, 0.000; *p* = 0.051). Greater support from friends was associated with higher baseline memory among men (β_between-effect_ = 0.173; 95% CI: 0.048, 0.299; *p* = 0.007) and among women (β_between-effect_ = 0.291; 95% CI: 0.168, 0.414; *p* < 0.001).

#### Sensitivity analysis

Estimates from the joint modelling of longitudinal and survival data showed a 13% reduced risk of death per unit increase in memory among both sexes (data not shown). In agreement with our main analysis, men with higher spousal strain showed faster decline in memory (β_between-effect×time_ = -0.025; 95% CI: -0.043, -0.007; *p* = 0.006). A similar but non-statistically significant finding was found for men with higher levels of strain from friends (β_between-effect×time_ = -0.017; 95% CI: -0.038, 0.003; *p* = 0.098), whilst men with higher levels of support from children showed slower decline (β_between-effect×time_ = 0.009; 95% CI: 0.000, 0.018; *p* = 0.051). A slower decline in memory was also observed among men with higher levels of spousal support (β_between-effect×time_ = 0.012; 95% CI: 0.004, 0.021; *p* = 0.003) (Tables S3-4).

## Discussion

Over a 16-year follow-up, we examined sex-specific longitudinal associations between the positive and negative dimensions of perceived relationship quality (social support and strain, respectively) and the rate of change in verbal memory among English community-dwelling persons aged 50-89 years at baseline. Our primary analyses showed that longitudinal associations were most prominent for men, varied by relationship type, and showed between-rather than within-person effects. Among men, a faster rate of decline in memory was associated with higher levels of strain from spouse and from friends; slower decline was associated with greater support from children.

### Comparisons with previous studies

Our findings agree to some extent with previous investigations of the social support/strain and cognitive ageing relationship. In the US Health and Retirement Study (HRS), higher strain with network members was associated with lower initial episodic memory, but not with the rate of change over 6-years (10). In the present study, higher social support (over all sources) was marginally associated with slower memory decline among men; this finding agrees with the study by Seeman et al (2001) which showed an association between greater frequency of emotionally supportive interactions with a network of social relations assessed at baseline and lower cognitive decline over a 7.5 year period (14).

Extending the follow-up period from 8-years in our previous investigation (27) to 16-years in our present study increased both statistical power and the ability to estimate the longer-term changes in verbal memory. In agreement with our previous analysis, differences in memory task performance were mainly influenced by between-persons differences in social support/strain, thereby confirming the utility of using REWB models to separate between- and within-person effects. Among men, both of our studies showed lower baseline memory and faster decline among those with higher levels of social strain from friends; among women, higher levels of strain from spouse and from children were associated with lower initial levels of memory but not with the rate of decline. Compared with the null findings in our previous investigation (27), our results presented herein for men showed higher levels of spousal strain and support from children to be associated with a faster and slower rate of decline, respectively. These new findings – which were robust to using a more complex model to account for potential selection bias due to informative dropout - potentially indicate that our previous analysis lacked sufficient power to identify these moderate associations between social strain and memory decline, which are now demonstrated via the longer follow-up. As such, our new findings potentially indicate the longer-term impacts of socially supportive/strained relations, suggesting the benefits of early interventions to enhance relationship quality.

### Potential mechanisms

Several psychological, behavioural and physiological pathways have been suggested in the literature for the associations between perceived relationship quality and cognitive ageing. Stress regulation (or the ‘stress-buffering’ hypothesis) could be a key mechanism, with supportive relations being a coping resource to reduce perceived stress, whilst strained relations act as a stressor (10, 11). Independently of social support, strained relations may associate more strongly with age-related cognitive decline due to patterns of physiological arousal, including increased risk for elevated inflammation (40). In addition to these pathways, the role of reverse causality cannot be eliminated, although we minimised its influence by excluding potentially cognitively unhealthy participants at baseline. Furthermore, previous studies have suggested that reverse causality is not an explanation through, for example, showing that initial cognitive performance did not predict subsequent changes in perceived relationship quality (16, 19).

### Implications for public health

Our study provides further evidence that differences between persons in mean levels of social support/strain associate with both initial levels of memory and with decline, even after adjustment for socio-demographics, healthy lifestyle behaviours, and physical- and mental-health. While the magnitude of the associations shown in our present study were modest, even minor differences in levels of cognitive functioning can over a period of several years substantially increase dependence and risk of adverse outcomes such as dementia (41). Levels of social support/strain and cognitive functioning are modifiable aspects of healthy ageing: therefore, it is imperative from a public health perspective to identify, design and implement interventions for middle and older-aged adults and their social network members in order to maximise the protective role that high-quality social relations can play in preserving cognitive functioning. Boosting social support levels through engagement in digital media is one possible intervention area worthy of investigation (42, 43). More specifically, our findings point to the importance of targeting interventions among men with higher than average levels of strained relations with their spouse and friends. Rather than addressing social relations in isolation, such strategies need to be placed within the context of multidomain interventions (44), which simultaneously target multiple risk-reducing psychosocial and lifestyle factors.

### Strengths and limitations

Strengths of our study include the benefits of analysing the ELSA cohort which includes its relatively large sample size (enabling sex-stratified analyses), multiple and detailed measurements of social support/strain across relationship types (allowing a more nuanced analysis than previous studies as evaluations of relationship quality can differ by type) and validated tests of verbal memory. Our analyses were strengthened by a longer follow-up period relative to other studies, enabling longer-term assessment of change in cognitive task performance. Our study was also strengthened by the use of joint modelling of longitudinal and survival data to examine the impact of study drop-out on the robustness of our main findings.

Our findings however should be interpreted cautiously due to several limitations. First, the assessments of social support/strain were based on self-reports. Our estimates could be subject to differential response bias if those with higher word recall scores are more likely to positively bias their perceptions of social support/strain, thereby potentially upwardly biasing the magnitude of associations (15). Secondly, adjustment for a wide range of variables may have led to an underestimation of effect sizes since some of the impact of social support/strain on verbal memory may be mediated through factors such as health behaviours, physical-and mental-health, and depressive symptomology. Thirdly, the large number of tests by fitting sex-specific models separately by relationship type increased the probability of detecting false associations (Type 1 error). We also acknowledge that the magnitude of the associations reported in our study for a specific relationship type may be moderated by the levels of support/strain available from sources; future research should investigate, for example, whether differences in the rate of cognitive decline by levels of spousal strain are modified by levels of support from children, friends or other family members. Fourthly, study drop out is a limitation inherent to analyses of cognitive ageing. Whilst random effects models are robust under the assumption that data is missing at random, and we adjusted for practice effects, our findings should still be interpreted in the context of a cohort that was increasingly selective over time, with those most healthy and affluent being most likely to remain. Fifthly, information on death via mortality linkage is currently available in the public datasets only up to wave 6 (2010-11); thereafter we could not identify those lost to the study through death. Finally, as in all observation studies, our findings could have been influenced by additional confounders such as personality characteristics that were not available.

## Conclusion

In conclusion, although modest in magnitude, our findings provide robust evidence for the notion that higher and lower levels of social support and social strain, respectively, can reduce the rate of cognitive decline in middle- and older-age, especially among men, over 16 years. These findings can inform future research studies and intervention strategies designed to maximise the potential role of high-quality social relations in achieving healthy cognitive ageing.

## Data Availability

ELSA datasets, including the harmonized dataset created as part of the gateway of global ageing data to facilitate cross-national comparisons, are available for free upon registration to the UK Data Service (https://www.ukdataservice.ac.uk/).
Oldfield, Z., Rogers, N., Phelps, A., Blake, M., Steptoe A., Oskala, A., Marmot, M., Clemens S., Nazroo J., Banks J. (2020) English Longitudinal Study of Ageing: Waves 0-9, 1998-2019. [data collection]. 33rd Edition. UK Data Service. SN: 5050. http://doi.org/10.5255/UKDA-SN-5050-20.
Analytical syntax to enable replication of the results using the datasets archived at the UK Data Service is available at https://github.com/shauns11/social_relations_and_memory.

https://github.com/shauns11/social_relations_and_memory

## Acknowledgements

The authors thank the interviewers and nurses, the participants in the ELSA study, and colleagues at NatCen Social Research. We thank the original data creators, depositors, copyright holders, the funders of the Data Collections and the UK Data Archive for the use of ELSA data. The original data creators, depositors or copyright holders bear no responsibility for the current analysis or interpretation.

## Funding

ELSA was developed by a team of researchers based at University College London, the Institute for Fiscal Studies, and NatCen Social Research. Funding was provided by the National Institute on Aging (grants 2RO1AG7644-01A1 and 2RO1AG017644) and a consortium of UK government departments. The present study was unfunded. JL is supported by the National Natural Science Foundation of China (#71804201), and the Natural Science Foundation of Guangdong Province (#2018A0303130046, #2017A030310346).

## Conflict of interest

Both authors declare no financial relationships with any organizations that might have an interest in the submitted work, and no other relationships or activities that could appear to have influenced the proposed work.

## Consent and ethical approval

ELSA participants provided signed consent for taking part in the study and for linkage to mortality data, and ethical approval for each wave was granted by the London Multicentre Research Ethics Committee (MREC/01/2/91). This study is a secondary analysis of previously collected data; therefore, additional ethical approval was not required.

## Data sharing statement

ELSA datasets, including the harmonized dataset created as part of the gateway of global ageing data to facilitate cross-national comparisons, are available for free upon registration to the UK Data Service (https://www.ukdataservice.ac.uk/).

> Oldfield, Z., Rogers, N., Phelps, A., Blake, M., Steptoe A., Oskala, A., Marmot, M., Clemens S., Nazroo J., Banks J. (2020) *English Longitudinal Study of Ageing: Waves 0-9, 1998-2019*. [data collection]. 33^rd^ Edition. UK Data Service. SN: 5050. http://doi.org/10.5255/UKDA-SN-5050-20

Analytical syntax to enable replication of the results using the datasets archived at the UK Data Service is available at https://github.com/shauns11/social_relations_and_memory.

### Abbreviations

ELSA: English Longitudinal Study of Ageing
HRS: Health and Retirement study
REWB: random effects within-between models

## Notes

### Competing Interest Statement

The authors have declared no competing interest.

## References

1. Sabia S, Guéguen A, Marmot MG, Shipley MJ, Ankri J, Singh-Manoux A. Does cognition predict mortality in midlife? Results from the Whitehall II cohort study. Neurobiology of Aging. 2010;31(4):688–95.

2. Batty GD, Deary IJ, Zaninotto P. Association of cognitive function with cause-specific mortality in middle and older age: follow-up of participants in the english longitudinal study of ageing. American Journal of Epidemiology. 2016;183(3):183–90.

3. Cohen S, Wills TA. Stress, social support, and the buffering hypothesis. Psychological Bulletin. 1985;98(2):310.

4. Organization WH. Noncommunicable Diseases and Mental Health Cluster. Noncommunicable Disease Prevention and Health Promotion Department. Aging and Life Course. Active aging: a policy framework. Geneva; 2002. Active ageing a policy framework.

5. Liao J, Brunner EJ. Structural and functional measures of social relationships and quality of life among older adults: does chronic disease status matter? Quality of Life Research. 2016;25(1):153–64.

6. Hu B, Li L. The protective effects of informal care receipt against the progression of functional limitations among Chinese older people. The Journals of Gerontology: Series B. 2020;75(5):1030–41.

7. Antonucci TC, Ajrouch KJ, Birditt KS. The convoy model: Explaining social relations from a multidisciplinary perspective. The Gerontologist. 2014;54(1):82–92.

8. Sharifian N, Sol K, Zahodne LB, Antonucci TC. Social relationships and adaptation in later life. Reference Module in Neuroscience and Biobehavioral Psychology. 2022.

9. Amieva H, Stoykova R, Matharan F, Helmer C, Antonucci TC, Dartigues J-F. What aspects of social network are protective for dementia? Not the quantity but the quality of social interactions is protective up to 15 years later. Psychosomatic Medicine. 2010;72(9):905–11.

10. Zahodne LB, Ajrouch KJ, Sharifian N, Antonucci TC. Social relations and age-related change in memory. Psychology and Aging. 2019;34(6):751.

11. Kuiper JS, Zuidersma M, Zuidema SU, Burgerhof JG, Stolk RP, Oude Voshaar RC, et al. Social relationships and cognitive decline: a systematic review and meta-analysis of longitudinal cohort studies. International Journal of Epidemiology. 2016;45(4):1169–206.

12. Newsom JT, Nishishiba M, Morgan DL, Rook KS. The relative importance of three domains of positive and negative social exchanges: a longitudinal model with comparable measures. Psychology and Aging. 2003;18(4):746.

13. Rook KS. Parallels in the study of social support and social strain. Journal of Social and Clinical Psychology. 1990;9(1):118–32.

14. Seeman TE, Lusignolo TM, Albert M, Berkman L. Social relationships, social support, and patterns of cognitive aging in healthy, high-functioning older adults: MacArthur studies of successful aging. Health Psychology. 2001;20(4):243.

15. Seeman TE, Miller-Martinez DM, Stein Merkin S, Lachman ME, Tun PA, Karlamangla AS. Histories of Social Engagement and Adult Cognition: Midlife in the U.S. Study. The Journals of Gerontology: Series B. 2011;66B(Suppl_1):i141–i52.

16. Liao J, Head J, Kumari M, Stansfeld S, Kivimaki M, Singh-Manoux A, et al. Negative aspects of close relationships as risk factors for cognitive aging. American Journal of Epidemiology. 2014;180(11):1118–25.

17. Pillemer S, Ayers E, Holtzer R. Gender-stratified analyses reveal longitudinal associations between social support and cognitive decline in older men. Aging & Mental Health. 2019;23(10):1326–32.

18. Chen Y, Feeley TH. Social support, social strain, loneliness, and well-being among older adults: An analysis of the Health and Retirement Study. Journal of Social and Personal Relationships. 2014;31(2):141–61.

19. Luo M, Edelsbrunner PA, Siebert JS, Martin M, Aschwanden D. Longitudinal Within-Person Associations Between Quality of Social Relations, Structure of Social Relations, and Cognitive Functioning in Older Age. The Journals of Gerontology: Series B. 2021.

20. Coventry WL, Gillespie N, Heath AC, Martin N. Perceived social support in a large community sample. Social Psychiatry and Psychiatric Epidemiology. 2004;39(8):625–36.

21. Antonucci TC, Akiyama H. An examination of sex differences in social support among older men and women. Sex Roles. 1987;17(11-12):737–49.

22. Fuhrer R, Stansfeld SA. How gender affects patterns of social relations and their impact on health: a comparison of one or multiple sources of support from “close persons”. Social Science & Medicine. 2002;54(5):811–25.

23. Van Tilburg T, Van Groenou MB. Network and health changes among older Dutch adults. Journal of Social Issues. 2002;58(4):697–713.

24. Gurung RA, Taylor SE, Seeman TE. Accounting for changes in social support among married older adults: insights from the MacArthur Studies of Successful Aging. Psychology and Aging. 2003;18(3):487.

25. Sophia R-H, Skrondal A. Multilevel and longitudinal modeling using Stata: Stata Press; 2012.

26. Twisk JW, de Vente W. Hybrid models were found to be very elegant to disentangle longitudinal within-and between-subject relationships. Journal of Clinical Epidemiology. 2019;107:66–70.

27. Liao J, Scholes S. Association of social support and cognitive aging modified by sex and relationship type: A prospective investigation in the English longitudinal study of ageing. American Journal of Epidemiology. 2017;186(7):787–95.

28. Gottesman RF, Rawlings AM, Sharrett AR, Albert M, Alonso A, Bandeen-Roche K, et al. Impact of differential attrition on the association of education with cognitive change over 20 years of follow-up: the ARIC neurocognitive study. American Journal of Epidemiology. 2014;179(8):956–66.

29. Steptoe A, Breeze E, Banks J, Nazroo J. Cohort profile: the English longitudinal study of ageing. International Journal of Epidemiology. 2013;42(6):1640–8.

30. Zaninotto P, Batty GD, Allerhand M, Deary IJ. Cognitive function trajectories and their determinants in older people: 8 years of follow-up in the English Longitudinal Study of Ageing. J Epidemiol Community Health. 2018;72(8):685–94.

31. Baars M, Van Boxtel M, Dijkstra J, Visser P, Van Den Akker M, Verhey F, et al. Predictive value of mild cognitive impairment for dementia. Dementia and Geriatric Cognitive Disorders. 2009;27(2):173–81.

32. Yin J, Lassale C, Steptoe A, Cadar D. Exploring the bidirectional associations between loneliness and cognitive functioning over 10 years: the English longitudinal study of ageing. International Journal of Epidemiology. 2019;48(6):1937–48.

33. Radloff LS. The CES-D scale: A self-report depression scale for research in the general population. Applied Psychological Measurement. 1977;1(3):385–401.

34. Hoffman L, Stawski RS. Persons as contexts: Evaluating between-person and within-person effects in longitudinal analysis. Research in Human Development. 2009;6(2-3):97–120.

35. Vivot A, Power MC, Glymour MM, Mayeda ER, Benitez A, Spiro III A, et al. Jump, hop, or skip: modeling practice effects in studies of determinants of cognitive change in older adults. American Journal of Epidemiology. 2016;183(4):302–14.

36. Karlamangla AS, Miller-Martinez D, Aneshensel CS, Seeman TE, Wight RG, Chodosh J. Trajectories of cognitive function in late life in the United States: demographic and socioeconomic predictors. American Journal of Epidemiology. 2009;170(3):331–42.

37. Enders CK. Analyzing longitudinal data with missing values. Rehabilitation Psychology. 2011;56(4):267.

38. Sabia S, Elbaz A, Dugravot A, Head J, Shipley M, Hagger-Johnson G, et al. Impact of smoking on cognitive decline in early old age: the Whitehall II cohort study. Archives of General Psychiatry. 2012;69(6):627–35.

39. Raitanen J, Stenholm S, Tiainen K, Jylhä M, Nevalainen J. Longitudinal change in physical functioning and dropout due to death among the oldest old: a comparison of three methods of analysis. European Journal of Ageing. 2019:1–10.

40. Yang YC, Schorpp K, Harris KM. Social support, social strain and inflammation: Evidence from a national longitudinal study of US adults. Social Science & Medicine. 2014;107:124–35.

41. Edwards JD, Wadley V, Vance D, Wood K, Roenker D, Ball K. The impact of speed of processing training on cognitive and everyday performance. Aging & Mental Health. 2005;9(3):262–71.

42. Liao J, Xiao H-Y, Li X-Q, Sun S-H, Liu S-X, Yang Y-J, et al. A Social Group-Based Information-Motivation-Behavior Skill Intervention to Promote Acceptability and Adoption of Wearable Activity Trackers Among Middle-Aged and Older Adults: Cluster Randomized Controlled Trial. JMIR mHealth and uHealth. 2020;8(4):e14969.

43. Dodge HH, Zhu J, Mattek NC, Bowman M, Ybarra O, Wild KV, et al. Web-enabled conversational interactions as a method to improve cognitive functions: Results of a 6-week randomized controlled trial. Alzheimer’s & Dementia: Translational Research & Clinical Interventions. 2015;1(1):1–12.

44. Ngandu T, Lehtisalo J, Solomon A, Levälahti E, Ahtiluoto S, Antikainen R, et al. A 2 year multidomain intervention of diet, exercise, cognitive training, and vascular risk monitoring versus control to prevent cognitive decline in at-risk elderly people (FINGER): a randomised controlled trial. The Lancet. 2015;385(9984):2255–63.

