## Supplementary Data for "Social support, social strain and declines in verbal memory: 16-year follow-up of the English Longitudinal Study of Ageing cohort"

^*^Address correspondence to:

### Supplementary Material

| **Contents** |  | **Page** |
| --- | --- | --- |
| **Tables:** |  |  |
| **S1** | Unadjusted linear random effects model for baseline levels and decline in memory | 3 |
| **S2** | Results from REWB models of the between-persons and within-person associations between social support/strain and verbal memory (across all sources) | 4 |
| **S3** | Results from Joint models of the between-persons and within-person associations between social support/strain and verbal memory (across all sources) | 6 |
| **S4** | Results from Joint models of the between-persons and within-person associations between social support/strain and verbal memory by relationship type | 7 |
| **Figure** |  |  |
| S1 | Flowchart of analytical sample for main analysis. | 8 |

**TABLE S1**. Unadjusted linear random effects model for baseline levels and decline in memory

| **Parameter** | **β** | **95% CI** | ***P*-value** |  | **β** | **95% CI** | ***P*-value** |
| --- | --- | --- | --- | --- | --- | --- | --- |
|  | *Men (n = 4595)* | | |  | *Women (n = 5514)* | | |
| **Fixed part:** |  |  |  |  |  |  |  |
| Constant | 9.3 |  |  |  | 10.1 |  |  |
| Time-since-baseline, waves | 0.096 | (0.052, 0.139) | **<0.001** |  | 0.153 | (0.112, 0.194) | **<0.001** |
| Age-at-baseline, years | -0.145 | (-0.154, -0.136) | **<0.001** |  | -0.138 | (-0.146, -0.130) | **<0.001** |
| Time^2^ | -0.043 | (-0.049, -0.037) | **<0.001** |  | -0.050 | (-0.056, -0.045) | **<0.001** |
| Age^2^ | -0.001 | (-0.002, 0.000) | **0.008** |  | -0.003 | (-0.004, -0.002) | **<0.001** |
| Time × age | -0.015 | (-0.017, -0.013) | **<0.001** |  | -0.020 | (-0.022, -0.018) | **<0.001** |
| **Random part:** |  |  |  |  |  |  |  |
| Intercept variance | 4.8 | (4.5, 5.1) |  |  | 4.8 | (4.5, 5.1) |  |
| Slope variance | 0.1 | (0.0, 0.1) |  |  | 0.1 | (0.1, 0.1) |  |
| Intercept-by-slope covariance | -0.03 | (-0.08, 0.03) |  |  | 0.00 | (-0.05, 0.05) |  |

**β**: beta coefficient; CI: confidence interval.

Age at baseline (wave 1: 2002-03) centred at 65 years. Results in bold indicate *P* < 0.05. Verbal memory trajectories estimated

from these models are shown graphically in Figure 1.

**TABLE S2**. Results from REWB models of the between-persons and within-person associations between social support/strain and verbal memory (across all sources)

| **Parameter** | **β** | **95% CI** | ***P*-value** |  | **β** | **95% CI** | ***P*-value** |
| --- | --- | --- | --- | --- | --- | --- | --- |
|  | *Men (n=4595)* | | |  | *Women (n=5514)* | | |
| **Fixed part:** |  |  |  |  |  |  |  |
| Constant | 10.7 | (10.4, 11.1) |  |  | 11.5 | (11.1, 11.8) |  |
| Time-since-baseline, years | -0.211 | (-0.452, 0.029) | 0.085 |  | -0.123 | (-0.347, 0.100) | 0.280 |
| Age-at-baseline, years | **-0.129** | **(-0.139, -0.120)** | **<0.001** |  | **-0.122** | **(-0.132, -0.112)** | **<0.001** |
| Time^2^ | -0.019 | (-0.056, 0.018) | 0.321 |  | -0.024 | (-0.059, 0.011) | 0.180 |
| Age^2^ | **-0.001** | **(-0.002, 0.000)** | **0.038** |  | **-0.003** | **(-0.003, -0.002)** | **<0.001** |
| Age × time | **-0.009** | **(-0.015, -0.004)** | **<0.001** |  | **-0.011** | **(-0.016, -0.006)** | **<0.001** |
| Age^2^ × time | 0.000 | (0.000, 0.000) | 0.673 |  | 0.000 | (0.000, 0.000) | 0.564 |
| Age × time^2^ | **-0.001** | **(-0.002, 0.000)** | **0.049** |  | **-0.001** | **(-0.002, 0.000)** | **0.022** |
| Prior memory tests | **0.363** | **(0.123, 0.604)** | **0.003** |  | **0.372** | **(0.152, 0.592)** | **0.001** |
| Prior memory tests × time | -0.028 | (-0.066, 0.011) | 0.157 |  | -0.031 | (-0.067, 0.006) | 0.100 |
| **Social support:** |  |  |  |  |  |  |  |
| Between-effect | 0.070 | (-0.082, 0.221) | 0.367 |  | 0.007 | (-0.143, 0.158) | 0.925 |
| Between-effect × time | 0.030 | (-0.002, 0.061) | 0.067 |  | 0.021 | (-0.010, 0.052) | 0.187 |
| Within-effect | -0.005 | (-0.184, 0.174) | 0.954 |  | 0.056 | (-0.123, 0.236) | 0.534 |
| Within-effect × time | -0.027 | (-0.074, 0.021) | 0.269 |  | -0.025 | (-0.069, 0.019_ | 0.264 |
| **Social strain:** |  |  |  |  |  |  |  |
| Between-effect | **-0.441** | **(-0.703, -0.179)** | **0.001** |  | -0.479 | (-0.732, -0.225) | **<0.001** |
| Between-effect × time | -0.029 | (-0.083, 0.026) | 0.302 |  | 0.025 | (-0.025, 0.075) | 0.331 |
| Within-effect | -0.069 | (-0.296, 0.159) | 0.554 |  | 0.179 | (-0.011, 0.369) | 0.065 |
| Within-effect × time | 0.004 | (-0.058, 0.065) | 0.909 |  | -0.043 | (-0.093, 0.006) | 0.084 |
| **Socioeconomic position:** |  |  |  |  |  |  |  |
| Education (middle) | **-1.186** | **(-1.399, -0.973)** | **<0.001** |  | **-0.941** | **(-1.194, -0.687)** | **<0.001** |
| Education (low) | **-2.270** | **(-2.515, -2.025)** | **<0.001** |  | **-2.093** | **(-2.368, -1.818)** | **<0.001** |
| Wealth (2) | -0.193 | (-0.411, 0.026) | 0.084 |  | -0.127 | (-0.336, 0.082) | 0.234 |
| Wealth (3) | **-0.416** | **(-0.648, -0.184)** | **<0.001** |  | **-0.452** | **(-0.670, -0.234)** | **<0.001** |
| Wealth (4) | **-0.425** | **(-0.672, -0.178)** | **0.001** |  | **-0.601** | **(-0.829, -0.373)** | **<0.001** |
| Wealth (lowest) | **-0.794** | **(-1.048, -0.539)** | **<0.001** |  | **-0.982** | **(-1.221, -0.743)** | **<0.001** |
| Education (middle) × time | **0.045** | **(0.000, 0.089)** | **0.048** |  | 0.007 | (-0.045, 0.059) | 0.804 |
| Education (low) × time | **0.077** | **(0.023, 0.131)** | **0.005** |  | -0.013 | (-0.072, 0.046) | 0.668 |
| Wealth (2) × time | -0.026 | (-0.069, 0.018) | 0.249 |  | 0.001 | (-0.044, 0.047) | 0.958 |
| Wealth (3) × time | -0.026 | (-0.075, 0.023) | 0.303 |  | 0.003 | (-0.044, 0.050) | 0.903 |
| Wealth (4) × time | -0.084 | (-0.137, -0.030) | **0.002** |  | -0.001 | (-0.052, 0.051) | 0.972 |
| Wealth (lowest) × time | -0.015 | (-0.072, 0.042) | 0.608 |  | 0.003 | (-0.051, 0.058) | 0.905 |
| **Health behaviours:** |  |  |  |  |  |  |  |
| Current smoker | 0.104 | (-0.090, 0.299) | 0.292 |  | 0.123 | (-0.054, 0.300) | 0.173 |
| Ever drunk alcohol | **0.466** | **(0.218, 0.714)** | **<0.001** |  | **0.499** | **(0.324, 0.675)** | **<0.001** |
| Physically inactive | **-0.302** | **(-0.498, -0.105)** | **0.003** |  | **-0.311** | **(-0.468, -0.154)** | **<0.001** |
| Current smoker × time | -0.026 | (-0.080, 0.029) | 0.354 |  | **-0.077** | **(-0.131, -0.022)** | **0.006** |
| Ever drunk alcohol × time | -0.030 | (-0.082, 0.023) | 0.265 |  | -0.033 | (-0.074, 0.007) | 0.109 |
| Physically inactive × time | -0.013 | (-0.060, 0.033) | 0.577 |  | -0.005 | (-0.043, 0.033) | 0.798 |
| **Social participation** |  |  |  |  |  |  |  |
| Social participation | 0.035 | (-0.011, 0.082) | 0.138 |  | **0.055** | **(0.013, 0.096)** | **0.011** |
| Social participation × time | 0.007 | (-0.004, 0.019) | 0.221 |  | **0.012** | **(0.001, 0.023)** | **0.030** |
| **Mobility limitations** |  |  |  |  |  |  |  |
| ADL | **-0.088** | **(-0.169, -0.008)** | **0.031** |  | 0.031 | (-0.036, 0.097) | 0.370 |
| ADL × time | 0.000 | (-0.020, 0.020) | 0.979 |  | **-0.018** | **(-0.034, -0.002)** | **0.025** |
| **Depressive symptoms** |  |  |  |  |  |  |  |
| CESD | **-0.069** | **(-0.106, -0.031)** | **<0.001** |  | **-0.048** | **(-0.080, -0.017)** | **0.003** |
| CESD × time | -0.001 | (-0.011, 0.010) | 0.900 |  | -0.003 | (-0.011, 0.005) | 0.446 |
| **Random part:** |  |  |  |  |  |  |  |
| Intercept (SD) | 1.9 | (1.9, 2.0) |  |  | 2.0 | (1.9, 2.0) |  |
| Slope (SD) | 0.21 | (0.19, 0.24) |  |  | 0.26 | (0.24, 0.28) |  |
| Intercept-by-slope correlation | -0.07 | (-0.18, 0.04) |  |  | -0.09 | (-0.17, 0.00) |  |

**β**= beta coefficient**;** CI = confidence interval; REWB: random effects within-between model; SD = standard deviation.

Age at baseline (wave 1: 2002-03) centred at 65 years. Results in bold indicate *P* < 0.05.

**TABLE S3.** Results from Joint models of the between-persons and within-person associations between social support/strain and verbal memory (across all sources)

| **Parameter** | **β** | **95% CI** | ***P*-value** |  | **β** | **95% CI** | ***P*-value** |
| --- | --- | --- | --- | --- | --- | --- | --- |
|  | *Men (n = 4499)* | | |  | *Women (n = 5379)* | | |
| **Social support:** |  |  |  |  |  |  |  |
| Between-effect | -0.012 | -0.149, 0.125 | 0.865 |  | -0.088 | -0.221, 0.045 | 0.193 |
| Between-effect × time | **0.017** | **0.002, 0.032** | **0.025** |  | 0.014 | 0.000, 0.029 | 0.057 |
| Within-effect | -0.043 | -0.236, 0.150 | 0.661 |  | -0.024 | -0.207, 0.159 | 0.799 |
| Within-effect × time | 0.000 | -0.024, 0.023 | 0.976 |  | 0.008 | -0.015, 0.030 | 0.501 |
| **Social strain:** |  |  |  |  |  |  |  |
| Between-effect | **-0.449** | **-0.670, -0.229** | **<0.001** |  | **-0.389** | **-0.603, -0.175** | **<0.001** |
| Between-effect × time | -0.013 | -0.038, 0.012 | 0.316 |  | 0.003 | -0.021, 0.028 | 0.781 |
| Within-effect | -0.110 | -0.342, 0.123 | 0.355 |  | 0.070 | -0.150, 0.290 | 0.532 |
| Within-effect × time | 0.019 | -0.010, 0.049 | 0.204 |  | 0.012 | -0.015, 0.039 | 0.397 |

β: beta coefficient; CI: confidence interval

Model adjusted for time-since-baseline (origin 0); time^2^; age (centered at 65); age^2^; time×age; time×age^2^; number of prior verbal memory assessments; education; wealth; smoking status; ever drunk alcohol; physical inactivity; social participation; depression; and mobility limitations. Results in bold indicate *P* < 0.05. Shaded cells indicate significant associations in our main analysis (see **Table 2**).

**TABLE S4.** Results from Joint models of the between-persons and within-person associations between social support/strain and verbal memory by relationship type

| **Parameter** | **Spouse** | | | **Children** | | | **Extended family members** | | | **Friends** | | |
| --- | --- | --- | --- | --- | --- | --- | --- | --- | --- | --- | --- | --- |
|  | **β** | **95% CI** | **P-value** | **β** | **95% CI** | **P-value** | **β** | **95% CI** | **P-value** | **β** | **95% CI** | **P-value** |
|  | *Men (n=4486)* | | | | | | | | | | | |
| **Social support:** |  |  |  |  |  |  |  |  |  |  |  |  |
| Between-effect | -0.017 | -0.096, 0.062 | 0.667 | 0.052 | -0.029, 0.132 | 0.206 | **-0.089** | **-0.168, -0.009** | **0.028** | 0.048 | -0.045, 0.141 | 0.311 |
| Between-effect×time | **0.012** | **0.004, 0.021** | **0.003** | **0.009** | **0.000, 0.018** | **0.051** | 0.002 | -0.008, 0.011 | 0.737 | 0.003 | -0.008, 0.015 | 0.566 |
| Within-effect | -0.074 | -0.201, 0.052 | 0.250 | 0.042 | -0.103, 0.186 | 0.569 | 0.001 | -0.090, 0.092 | 0.983 | 0.034 | -0.070, 0.138 | 0.522 |
| Within-effect×time | 0.010 | -0.005, 0.025 | 0.209 | -0.002 | -0.019, 0.016 | 0.840 | -0.006 | -0.016, 0.005 | 0.288 | -0.004 | -0.016, 0.009 | 0.554 |
| **Social strain:** |  |  |  |  |  |  |  |  |  |  |  |  |
| Between-effect | -0.065 | -0.232, 0.103 | 0.449 | -0.119 | -0.274, 0.036 | 0.133 | **-0.384** | **-0.521, -0.248** | **<0.001** | **-0.392** | **-0.556, -0.228** | **<0.001** |
| Between-effect×time | **-0.025** | **-0.043, -0.007** | **0.006** | -0.001 | -0.019, 0.017 | 0.937 | -0.002 | -0.019, 0.015 | 0.829 | -0.017 | -0.038, 0.003 | 0.098 |
| Within-effect | -0.148 | -0.315, 0.020 | 0.084 | 0.021 | -0.133, 0.176 | 0.786 | 0.014 | -0.109, 0.137 | 0.820 | -0.066 | -0.209, 0.076 | 0.362 |
| Within-effect×time | 0.007 | -0.014, 0.028 | 0.499 | 0.003 | -0.016, 0.022 | 0.773 | 0.005 | -0.011, 0.020 | 0.561 | 0.013 | -0.005, 0.031 | 0.157 |
|  | *Women (n=5356)* | | | | | | | | | | | |
| **Social support:** |  |  |  |  |  |  |  |  |  |  |  |  |
| Between-effect | **-0.092** | **-0.165, -0.019** | **0.013** | 0.037 | -0.039, 0.113 | 0.343 | -0.047 | -0.122, 0.028 | 0.219 | **0.210** | **0.118, 0.302** | **<0.001** |
| Between-effect×time | 0.007 | -0.001, 0.016 | 0.091 | 0.000 | -0.008, 0.008 | 0.984 | 0.005 | -0.004, 0.014 | 0.268 | -0.002 | -0.013, 0.009 | 0.741 |
| Within-effect | -0.024 | -0.131, 0.082 | 0.657 | 0.078 | -0.072, 0.228 | 0.308 | -0.073 | -0.156, 0.011 | 0.090 | -0.031 | -0.133, 0.071 | 0.546 |
| Within-effect×time | 0.006 | -0.006, 0.019 | 0.331 | -0.012 | -0.031, 0.006 | 0.178 | 0.002 | -0.008, 0.012 | 0.669 | **0.013** | **0.001, 0.025** | **0.034** |
| **Social strain:** |  |  |  |  |  |  |  |  |  |  |  |  |
| Between-effect | -0.136 | -0.286, 0.013 | 0.074 | **-0.247** | **-0.395, -0.098** | **0.001** | -0.108 | -0.237, 0.020 | 0.097 | -0.070 | -0.230, 0.090 | 0.393 |
| Between-effect×time | 0.003 | -0.014, 0.021 | 0.691 | 0.004 | -0.013, 0.021 | 0.656 | -0.003 | -0.019, 0.012 | 0.674 | -0.005 | -0.025, 0.015 | 0.626 |
| Within-effect | 0.081 | -0.069, 0.230 | 0.290 | 0.089 | -0.059, 0.237 | 0.239 | -0.039 | -0.151, 0.074 | 0.502 | -0.035 | -0.169, 0.100 | 0.614 |
| Within-effect×time | -0.005 | -0.023, 0.013 | 0.586 | -0.007 | -0.025, 0.011 | 0.445 | 0.013 | -0.001, 0.026 | 0.066 | 0.011 | -0.005, 0.028 | 0.177 |

β: beta coefficient; CI: confidence interval

Model adjusted for time-since-baseline (origin 0); time^2^; age (centered at 65); age^2^; time×age; time×age^2^; number of prior verbal memory assessments; education; wealth; smoking status; ever drunk alcohol; physical inactivity; social participation; depression; and mobility limitations. Results in bold indicate *P* < 0.05. Shaded cells indicate significant associations in our main analysis (see **Table 3**).

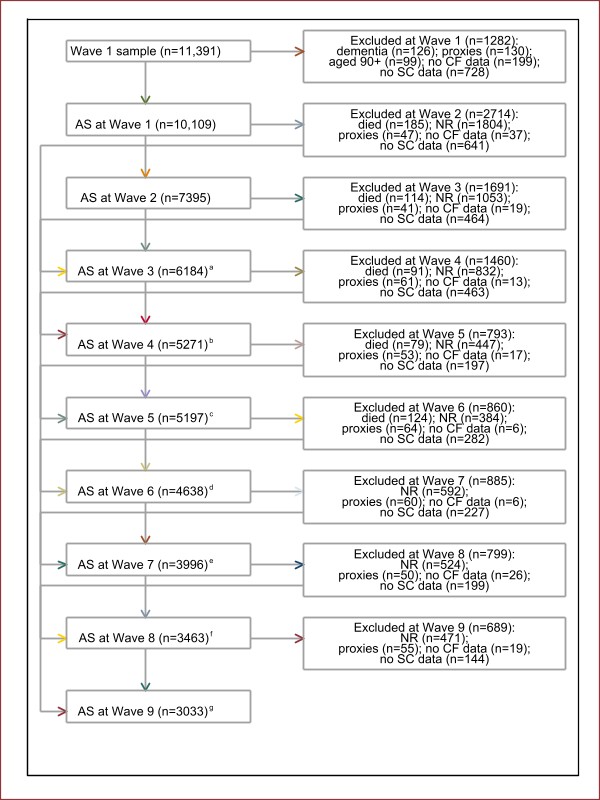

**FIGURE S1.** Flowchart of analytical sample for main analysis.

Selection of analytical sample (verbal memory data and filled-in self-completion questionnaires). The analytical sample comprised *n* = 10,109 participants aged 50-89 years at baseline (wave 1: 2002-03); whom contributed 49,286 observations over the study period (nine waves). Participants who missed a particular wave (e.g. through wave non-response) were allowed to reenter the analytical sample at future waves:

^a^ Includes n=480 not in the AS at Wave 2; ^b^ includes n=547 not in the AS at Wave 3; ^c^ includes n=719 not in the AS at Wave 4; ^d^ includes n=301 not in the AS at Wave 5; ^e^ includes n=243 not in the AS at Wave 6; ^f^ includes n=266 not in the AS at Wave 7; ^g^ includes n=259 not in the AS at Wave 8.

AS: analytical sample; CF: cognitive function; NR: non-response; SC: self-completion.
